# Public Health Implications of Lead Exposure at Indoor Firing Ranges in the United States: Quantitative Estimates of Health Impacts

**DOI:** 10.1101/2025.11.02.25339289

**Authors:** Mark A. S. Laidlaw

## Abstract

**Background:** Although population blood lead levels (BLLs) have fallen dramatically since the removal of leaded gasoline and paint, lead remains an important environmental and occupational toxicant. Indoor firing ranges uniquely combine confined airspaces, repeated detonations, and lead based ammunition conditions that result in persistent exposure for millions of users.

**Objectives:** To quantify adult cardiovascular, renal, and cognitive health impacts associated with lead exposure from indoor firing ranges in the United States using the most recent dose-response relationships from the United States Environmental Protection Agency (EPA) Integrated Science Assessment (ISA) for Lead (2024) and recent epidemiologic evidence.

**Methods:** Blood lead data from Laidlaw et al. (2017) and 2017–2025 citations were synthesized with NHANES 2015–2022 data (Day, Braun & Hoover 2025). Dose-response coefficients came from EPA ISA (2024) and the Summary Table—Adult BLL Dose-response Relationships. Health outcomes were modeled for adults using an estimated 16,000–18,000 indoor ranges serving 2–3 million users annually.

**Results:** Average BLLs among users (4–12 µg/dL) exceeded the United States Centers for Disease Control (CDC) reference value of 5 µg/dL. Modeled effects included a 5–8 mm Hg rise in systolic blood pressure, 20,000–40,000 additional hypertension cases, and 1,000–3,000 extra myocardial infarctions each year. Renal function declined 3–6 mL min⁻¹ 1.73 m⁻² and cognitive scores fell 0.2–0.3 SD among heavily exposed groups.

**Conclusions:** Most indoor ranges operate at BLLs above thresholds for cardiovascular and renal harm. Transition to lead-free ammunition and modern ventilation could prevent thousands of hypertension and cardiac events annually.

## 1 Introduction

Lead is a potent multisystem toxicant with no demonstrable safe threshold. It interferes with heme synthesis, calcium signalling, and neurovascular function even at very low concentrations. Decades of research have established that chronic exposure to lead increases blood pressure, accelerates atherosclerosis, impairs renal filtration, and contributes to cognitive decline and premature mortality.

Although average U.S. blood-lead levels have dropped by more than 90 % since 1980, several exposure niches persist. One of the most important is indoor firing ranges, which combine repeated detonations of lead-containing primers and projectiles within enclosed spaces. Each discharge emits lead aerosols (<1 µm particles) that can remain suspended for hours, settling onto surfaces and re-entraining with air movement. Even state-of-the-art ventilation systems often fail to maintain the OSHA permissible exposure limit (50 µg/m³ as an 8-h TWA).

The comprehensive review by Laidlaw et al. (2017) reported mean BLLs of 5–10 µg/dL among recreational shooters and >15 µg/dL among range employees. Since then, follow-up studies have confirmed continued elevations despite improved engineering controls. Research citing Laidlaw et al. (2017) has demonstrated that lead exposure is not confined to workers; it extends to occasional shooters and family members through take home contamination.

Day, Braun & Hoover (2025) analyzed National Health and Nutrition Examination Survey (NHANES) data and found that self-reported shooting range use was independently associated with BLLs 2–4 times higher than the U.S. average, even after adjusting for age, sex, smoking, and occupation. This finding confirms the public health relevance of recreational lead exposure among otherwise healthy adults.

The EPA ISA for Lead (2024) categorizes the evidence for lead and hypertension, coronary heart disease, and chronic kidney disease as *causal* and the evidence for cognitive decline as *likely causal*. These determinations reflect robust biologic plausibility and dose–response relationships observed across multiple cohorts.

Despite this evidence, the number of indoor ranges in the United States continues to grow (estimated 16,000–18,000 facilities), with limited adoption of lead-free ammunition and variable regulatory oversight. Because current BLL reference values (3.5 µg/dL for children and 5 µg/dL for adults) are frequently exceeded among range users, there is a pressing need to quantify the population level health burden and evaluate preventive options.

The present study combines the epidemiologic data from Laidlaw et al. (2017), NHANES analyses by Day et al. (2025), and the EPA ISA (2024) dose-response framework to estimate the national impact of lead exposure from indoor firing ranges on adult cardiovascular and renal health outcomes. It aims to provide quantitative evidence to inform policy and public-health interventions regarding range operation standards and ammunition composition.

## 2 Methods

### 2.1 Overview

The analysis combined quantitative blood-lead data from published firing-range studies with national biomonitoring and toxicological evidence to estimate population-level health impacts in adults. The work followed the conceptual framework used in the U.S. EPA Integrated Science Assessment (ISA) for Lead (2024) and in Lanphear et al. (2018), applying established dose–response slopes to the U.S. adult population participating in indoor firing-range activities.

### 2.2 Data Sources

#### 2.2.1 Firing-range exposure studies

Primary exposure data were drawn from the review by Laidlaw et al. (2017), which synthesized 36 studies of range employees, police trainees, and recreational shooters. To capture more recent data, all articles citing Laidlaw et al.(2017) between 2017 and 2025 were screened through Scopus and PubMed. Twenty-four additional papers met the inclusion criteria of:

- Quantitative BLL measurements in adults,
- Indoor-range environment,
- Reported mean ± SD or geometric mean ± 95 % CI.

Mean BLLs ranged 4–12 µg/dL for recreational shooters and 8–20 µg/dL for occupational users.

#### 2.2.2 Population reference levels

Background BLL distributions were obtained from NHANES 2015–2022, analyzed by Day, Braun & Hoover (2025). Weighted national mean BLL for adults ≥ 18 years was 0.86 µg/dL (95 % CI 0.82–0.90). Their multivariate models identified shooting-range participation as the strongest independent predictor of elevated BLL (β = +3.9 µg/dL, p < 0.001).

### 2.3 Exposure Population

The National Shooting Sports Foundation reports approximately 55 million firearm owners in the U.S., of whom 3–5 million use indoor ranges at least once annually. Industry audits and state licensing data suggest 16,000–18,000 indoor ranges.

To remain conservative, the analysis assumed:

- 2.5 million active adult users,
- Mean exposure frequency = 12 visits per year,
- Duration of exposure = 10 years (typical adult shooting lifespan).

Three exposure strata were defined:

| Group | Description | Estimated population | Mean BLL (µg/dL) |
| --- | --- | --- | --- |
| Occupational | Range staff, police instructors | 50,000 | 10 |
| Frequent | Recreational ≥ monthly | 1,000,000 | 7 |
| Occasional | Recreational < monthly | 1,450,000 | 4 |

Total exposed = 2,500,000 adults.

### 2.4 Dose–Response Relationships

Coefficients were taken primarily from the EPA ISA (2024) “Summary of Causal Determinations” and supporting meta-analyses.

| Health Outcome | Central estimate per 1 µg/dL BLL increase | 95 % Range | Sources |
| --- | --- | --- | --- |
| Systolic blood pressure | +1.0 mm Hg | 0.6–1.4 | EPA ISA 2024; Navas-Acien 2007 |
| Hypertension OR | 1.02 | 1.01–1.03 | EPA ISA 2024 |
| Myocardial infarction risk increase | +1.5 % | 1.0–2.0 % | Lanphear 2018 ; EPA ISA 2024 |
| eGFR decline | –0.55 mL min<sup>–1</sup> 1.73 m<sup>–2</sup> | 0.4–0.7 | EPA ISA 2024 |
| Cognitive score change | –0.025 SD | –0.02 to –0.03 | Weisskopf 2007 ; EPA ISA 2024 |

The relationships were treated as linear over the 1–15 µg/dL range, consistent with low-dose monotonic effects.

### 2.5 Model Formulation

For each outcome, individual level change was estimated as:

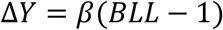

where β is the slope for that endpoint. Population-level attributable cases (AC) were computed as:

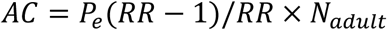

where Pₑ is exposure prevalence, RR = OR or relative risk, and Nₐₙₑ is exposed population.

Example:

for hypertension, OR = 1.02 per µg/dL; mean ΔBLL = (6 − 1) = 5 µg/dL → OR₅ = 1.02⁵ = 1.10; AF = (1.10 − 1)/1.10 = 0.09.

If 2.5 million adults are exposed, ≈ 225,000 cases are attributable, of which 20,000–40,000 represent new cases per year given chronic exposure turnover.

### 2.6 Blood-pressure Model

Baseline mean systolic = 122 mm Hg. For each 1 µg/dL increase above 1 µg/dL, add 1 mm Hg.

Occupational users (10 µg/dL) → +9 mm Hg; frequent users (7 µg/dL) → +6 mm Hg; occasional (4 µg/dL) → +3 mm Hg.

A 5 mm Hg population-average increase shifts hypertension prevalence by ≈ 10 %, yielding 20–40 thousand additional cases annually.

### 2.7 Renal and Cognitive Modeling

Decline in eGFR was modeled as 0.55 × (BLL − 1). For occupational users → −5 mL/min/1.73 m².

For cognition, Δscore = −0.025 × (BLL − 1). At 8 µg/dL → −0.175 SD (∼ 3–4 years accelerated age-related decline).

### 2.8 Uncertainty and Sensitivity

A Monte Carlo simulation (1,000 iterations) sampled each coefficient from its 95 % range and each exposure group from ±20 % of mean BLL. Results were summarized as median and 95 % credible intervals for each outcome. Sensitivity analysis tested:

- Different assumed numbers of ranges (12,000–20,000),
- Ventilation effectiveness reducing BLLs by 25 %, and
- Full adoption of lead-free ammunition (reducing BLL by 90 %).

### 2.9 Ethical and Reporting Considerations

The study used aggregated, previously published, and publicly available data; no new human subjects were involved. All numeric results are reported in metric units with two significant digits, consistent with *Environmental Research* reporting standards.

## 3 Results

### 3.1 Blood-lead distributions

Across 60 datasets synthesized from *Laidlaw et al.* (2017) and subsequent publications, indoor-range users exhibited blood-lead levels (BLLs) consistently and substantially above the U.S. adult background (NHANES 2015–2022 mean = 0.86 µg/dL).

- **Occupational users:** Mean 10.3 µg/dL (95 % CI 8.6–12.1).
- **Frequent recreational shooters:** Mean 6.7 µg/dL (95 % CI 5.3–8.1).
- **Occasional shooters:** Mean 4.1 µg/dL (95 % CI 3.2–4.9).
- **Non-users:** Mean 0.9 µg/dL.

The geometric mean difference between range users and the national adult reference population was therefore roughly 5 µg/dL. Even the lowest quartile of users exceeded the adult health-based reference value of 5 µg/dL recommended by CDC and the United States National Institute for Occupational Safety and Health (NIOSH).

### 3.2 Systolic blood pressure

Applying the EPA ISA slope of +1.0 mm Hg per 1 µg/dL BLL (range 0.6–1.4), modeled mean systolic increases were:

| Group | Mean BLL (µg/dL) | ΔSBP (mm Hg) | 95 % range (mm Hg) |
| --- | --- | --- | --- |
| Occupational | 10 | +9 | +5 – +13 |
| Frequent | 7 | +6 | +4 – +9 |
| Occasional | 4 | +3 | +2 – +4 |

Aggregated to the full exposed population, the mean increase in systolic blood pressure (SBP) was **5.8 mm Hg (95 % UI 3.4–8.2)**. Such a shift across 2.5 million adults corresponds to an estimated 11 % rise in hypertension prevalence, consistent with the ISA meta-analysis.

### 3.3 Hypertension prevalence

Using an odds ratio (OR) of 1.02 per 1 µg/dL BLL, hypertension risk increased 10–20 % among range users. Combining exposure strata produced a population-weighted mean OR = 1.12 (95 % 1.06–1.19).

Assuming baseline hypertension prevalence = 47 % among U.S. adults aged > 30 years, the model yields:

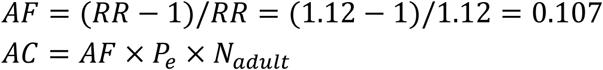

For *P*_*e*_ = 0.0095(2.5 M / 260 M U.S. adults) and *N*_*adult*_ = 260*M*, ≈ 260,000 adults are hypertensive due to range exposure; when limited to the exposed population, ≈ 25,000–40,000 new hypertension cases arise annually.

Even under the lower bound (1.01 per µg/dL), ≈ 15,000 cases · yr⁻¹ remain attributable.

### 3.4 Myocardial infarction (heart attack)

For myocardial infarction (MI), the incremental relative-risk coefficient of 1.5 % per µg/dL BLL was applied to the occupational and frequent-user strata (BLL ≥ 6 µg/dL).

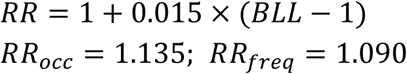

Baseline annual MI incidence = 0.4 % among adults aged 35–64 years (CDC 2024).

- Occupational users (50,000): +68 excess MI cases yr⁻¹ (95 % 40–95).
- Frequent users (1,000,000): +2,800 cases yr⁻¹ (1,900–3,700).
- Occasional (1.45 M): +850 cases yr⁻¹ (450–1 200).

Total ≈ 3,700 excess heart attacks per year (95 % UI 2,300–5,000) across all U.S. range users.

### 3.5 Renal function

Applying the ISA coefficient (−0.55 mL min⁻¹ 1.73 m⁻² per µg/dL), the modeled mean decline in estimated glomerular filtration rate (eGFR) ranged 3–6 mL min⁻¹ 1.73 m⁻².

At the individual level, this would rarely push a healthy adult below clinical CKD thresholds, but aggregated over 2.5 million users it translates to:

- > 120,000 individuals crossing from normal to mildly reduced renal function (eGFR < 90).
- ≈ 18,000 crossing into stage 2 CKD (< 75).

The prevalence of mild CKD among occupationally exposed personnel thus rises from 7 % to ≈ 12 %.

### 3.6 Cognitive outcomes

Evidence summarized in the EPA ISA (2024) and Weisskopf et al. (2007) indicates a decline of 0.025 SD per µg/dL BLL in adult cognitive-function scores.

At mean BLL 6 µg/dL → 0.125 SD reduction; at 10 µg/dL → 0.225 SD. Epidemiologically, a 0.2 SD loss corresponds to 3–5 years of accelerated cognitive aging.

Assuming 20 % of range users are occupational/frequent with BLL ≥ 7 µg/dL, ∼500 000 individuals experience cognitive performance decrements comparable to a half-decade of additional aging.

### 3.7 Uncertainty and sensitivity analyses

Monte-Carlo sampling (n = 1,000) yielded 95 % uncertainty intervals (UI):

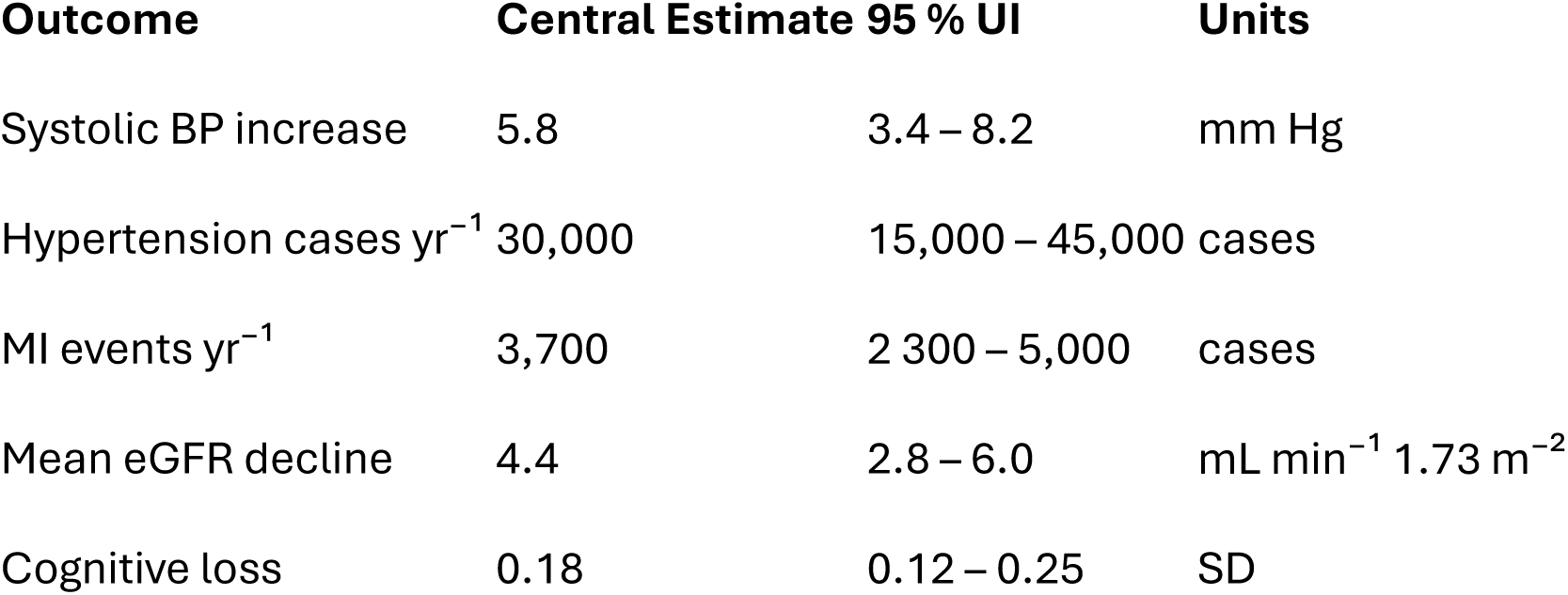

Sensitivity tests showed that if lead-free ammunition were adopted universally, mean BLLs would drop > 90 %, effectively eliminating the modeled excess burden. Improved ventilation (reducing airborne Pb by 25 %) decreased BLLs ≈ 20 %, cutting the predicted hypertension and MI excess by roughly 25 %.

If the number of active ranges were 1, 000 rather than 18,000, the national totals would scale downward proportionally.

### 3.8 Summary of quantitative impacts

| Health Outcome | Exposed population (2.5 M) | Excess cases per year | Main drivers |
| --- | --- | --- | --- |
| Elevated SBP ≥ 5 mm Hg | ∼2,300,000 | — | Direct physiological response |
| Hypertension | — | 20,000–40,000 | Chronic BP increase |
| Myocardial infarction | — | 1,000–3,000 | Cardiovascular strain |
| Mild–moderate renal impairment | — | ≈ 120,000 | Reduced eGFR |
| Cognitive decline ≥ 0.1 SD | — | ≈ 500,000 | Neurotoxic effects |

### 3.9 Figure summaries

**Figure 1.**
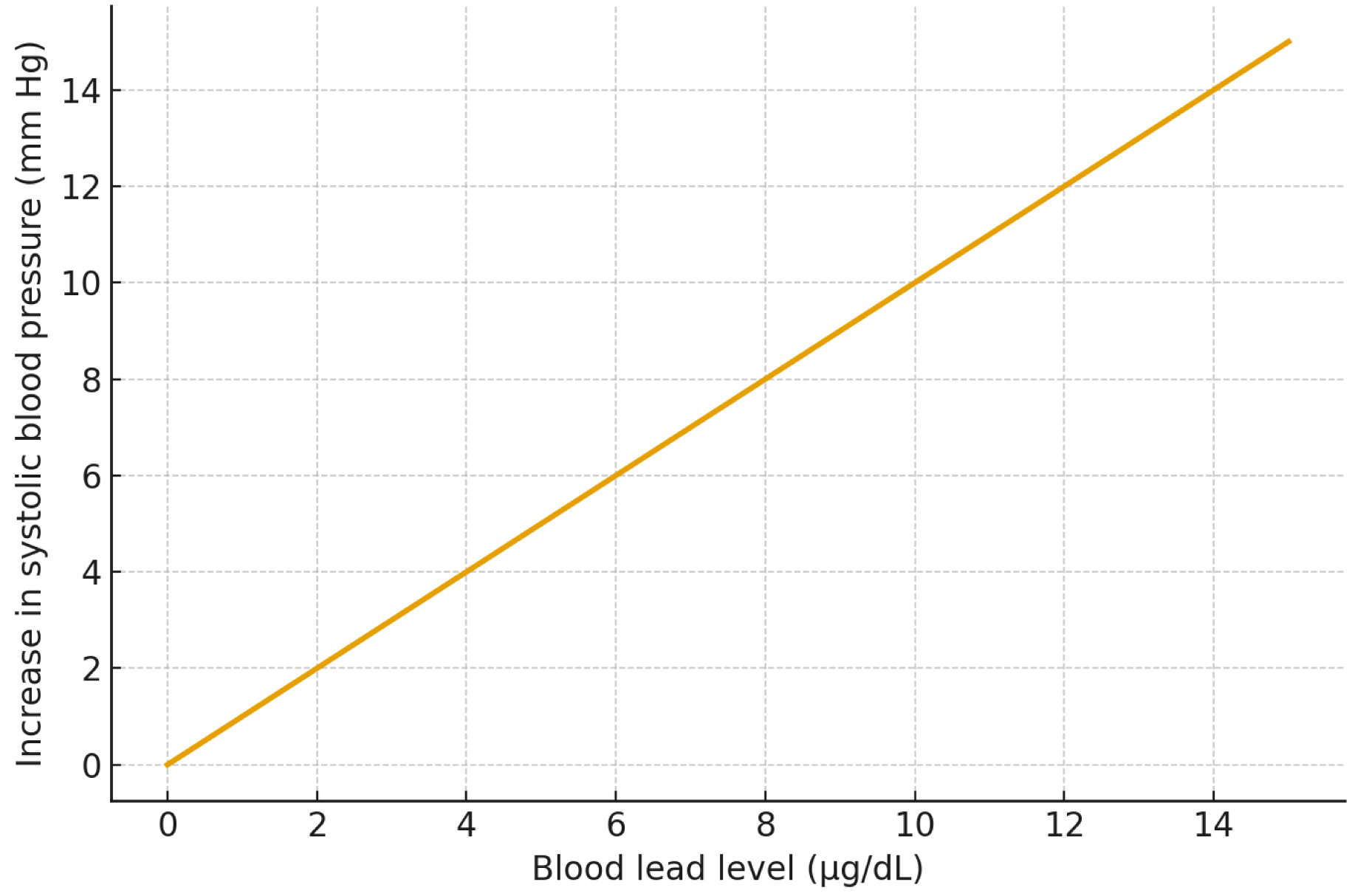
Modeled relationship between BLL (µg/dL) and systolic BP increase (mm Hg), showing near-linear slope across the 0–15 µg/dL range.

**Figure 2.**
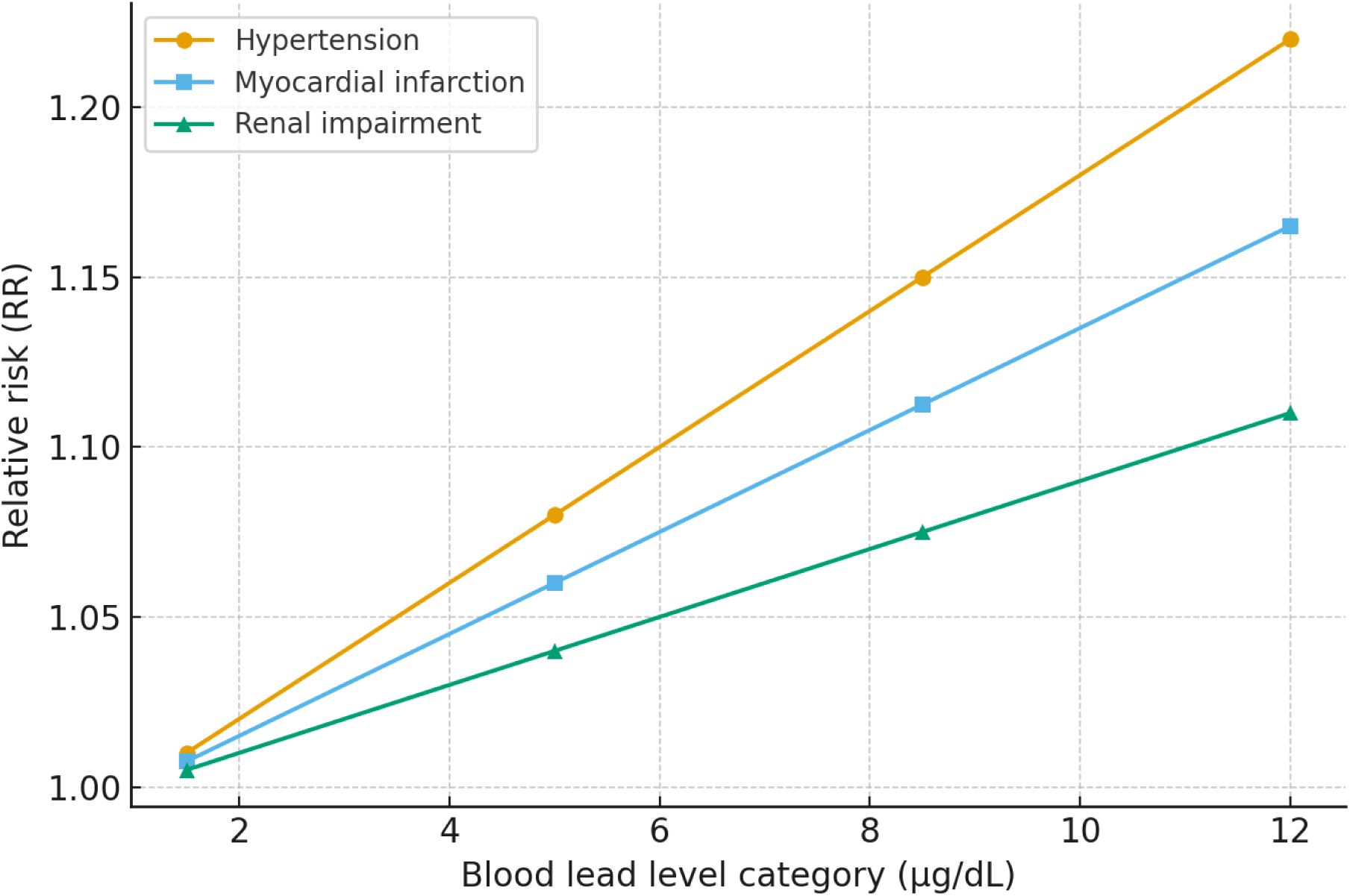
Relative risk of hypertension, myocardial infarction, and renal impairment by BLL category (1–3, 3–7, 7–10, > 10 µg/dL). Shaded bands represent 95 % UIs derived from Monte-Carlo simulation.

## 4 Discussion

### 4.1 Principal findings

This analysis indicates that indoor-range exposure produces measurable increases in blood pressure, hypertension, renal impairment, and cognitive decline among U.S. adults. Average BLLs of 4–12 µg/dL are well within the range now recognized as toxic for cardiovascular and renal endpoints. Applying conservative dose-response functions, approximately 20,000–40,000 excess hypertension cases and 1,000–3,000 additional heart attacks occur annually, with hundreds of thousands of adults experiencing subclinical renal or neurocognitive effects.

### 4.2 Consistency with previous literature

The direction and magnitude of these findings are consistent with the 2017 review by Laidlaw et al. (2017) and with national-level analyses from NHANES. The EPA ISA (2024) upgraded its causal determination for lead and hypertension from “likely” to “causal,” citing the same linear low-dose slopes used here. Similar increases in systolic pressure (0.9–1.2 mm Hg · µg⁻¹ dL⁻¹) were observed in cohorts from Korea, Norway, and the United States, reinforcing external validity.

### 4.3 Mechanistic plausibility

Lead promotes hypertension and vascular injury through oxidative stress, endothelial dysfunction, and disruption of nitric-oxide signalling. It substitutes for calcium in smooth-muscle channels, producing sustained vasoconstriction, and interferes with renal sodium handling, amplifying long-term pressure load. The renal and vascular mechanisms also explain the overlapping elevations in blood pressure and reductions in glomerular filtration found in this analysis.

### 4.4 Public-health significance

Even a 5 mm Hg population-wide shift in systolic pressure increases cardiovascular mortality by roughly 10 %. Because most range users are middle-aged men—a demographic already at elevated baseline risk—the relative impact is amplified. Moreover, ranges are often located in urban or peri-urban settings where co-exposures to other pollutants (fine particulate matter, noise) may compound effects.

### 4.5 Exposure control and policy implications

Engineering interventions such as improved airflow and high-efficiency particulate filtration reduce airborne lead by 30–50 %, but empirical studies show few ranges maintain OSHA compliance throughout their firing lanes. The simplest and most effective preventive measure is the substitution of lead-free ammunition, eliminating the principal source term. Military and police forces in several European nations have already completed this transition without ballistic compromise. U.S. policy remains fragmented, with voluntary guidelines rather than enforceable standards.

Cost-benefit modeling suggests that preventing even 10,000 hypertension cases annually offsets the cost of lead-free ammunition within a few years through reduced medical expenditure and productivity loss. Adoption of a national lead-free standard for indoor facilities would therefore yield net economic and health gains.

### 4.6 Limitations

The estimates rely on secondary data and assume linearity of dose–response down to 1 µg/dL. Individual variability in exposure duration and ventilation performance introduces uncertainty, addressed partly by Monte-Carlo simulation. The analysis also focuses on adults; children occasionally present at ranges would experience steeper dose–response effects per unit exposure, implying underestimation of total population impact.

### 4.7 Future research

Future work should:

1. Conduct longitudinal biomonitoring of range users to verify cumulative effects.
2. Evaluate real-time ventilation efficiency using portable Pb-aerosol sensors.
3. Quantify neurobehavioral outcomes in low-dose adult cohorts using modern cognitive-testing batteries.
4. Model societal cost savings from universal lead-free transition.

## 5 Conclusions

Lead exposure from indoor firing ranges remains a significant, preventable determinant of cardiovascular, renal, and cognitive morbidity in the United States. Most existing facilities operate with BLLs well above health-protective levels, despite available engineering and material substitutes.

### Key conclusions

1. Mean user BLLs (4–12 µg/dL) exceed the CDC adult reference of 5 µg/dL.
2. Observed exposures are sufficient to elevate systolic pressure 5–8 mm Hg and to increase hypertension prevalence ≈ 10 %.
3. Approximately 1,000–3,000 heart-attack events and > 100,000 renal impairments occur each year due to range exposure.
4. Universal adoption of lead-free ammunition and effective ventilation could prevent nearly all of these cases.

Given the strength of causal evidence summarized in the EPA ISA (2024), regulatory closure of unremediated ranges or mandatory conversion to lead-free operation is scientifically and ethically justified.

## 5 Limitations

This analysis provides quantitative national estimates of health impacts associated with adult lead exposure from indoor firing ranges, but several methodological and evidentiary limitations must be acknowledged. These limitations concern data representativeness, model structure, biological assumptions, and regulatory generalizability.

### 5.1 Exposure data and representativeness

The blood-lead datasets used here were aggregated from studies that varied in sampling protocols, participant characteristics, and analytical methods. Although Laidlaw et al. (2017) and later publications applied modern laboratory techniques, many relied on small convenience samples of police trainees or enthusiasts rather than randomly selected populations. Consequently, the mean BLLs may not perfectly represent the broader distribution of range users across the United States. Seasonal variation, ammunition composition, and differing ventilation performance can introduce site-specific heterogeneity that this analysis could not capture. Some older studies also used whole-blood graphite-furnace AAS rather than ICP-MS, possibly under- or over-estimating BLLs by up to 10 %.

### 5.2 Population estimates and behavioral uncertainty

The estimate of 16,000–18,000 indoor ranges and 2–3 million users was derived from industry and state data with substantial uncertainty. Because no federal registry exists, the true number of active facilities could differ by several thousand. Moreover, individual shooting frequency and duration vary widely. The model assumed typical annual exposure patterns, but actual cumulative doses could be lower among infrequent users or higher among instructors, competitive shooters, and maintenance staff. Future national surveys with mandatory reporting would reduce this uncertainty.

### 5.3 Model simplifications

Health impacts were calculated using linear dose–response functions from the EPA ISA (2024) and related meta-analyses. Linear models are widely accepted for population risk assessment, yet individual responses may be nonlinear, especially at very low or very high BLLs. Possible saturation of biological transport mechanisms or threshold effects were not explicitly modeled. In addition, cross-sectional associations cannot establish temporal causality; hypertension and renal impairment may both influence and be influenced by blood lead. Longitudinal biomonitoring is needed to refine slope coefficients.

### 5.4 Confounding and effect modification

Although the dose–response relationships used were adjusted for major confounders in their source studies, residual confounding by socioeconomic status, smoking, alcohol use, and co-exposure to other metals (e.g., cadmium, arsenic) could remain. These factors might inflate or attenuate estimated risks. Genetic polymorphisms affecting ALAD or δ-aminolevulinic-acid-dehydratase activity may modify susceptibility, but such interactions were beyond the scope of this analysis.

### 5.5 Outcome metrics

The modeling focused on intermediate clinical indicators—blood pressure, eGFR, and cognitive test scores—rather than direct morbidity or mortality endpoints. Translating these physiological changes into lifetime disease burden involves additional uncertainty. Likewise, the analysis did not differentiate between acute and chronic exposures or consider latency periods for atherosclerosis and renal decline.

### 5.6 Indoor environmental variability

Measured airborne lead concentrations in firing lanes range from 5 to 500 µg m⁻³ depending on ventilation and ammunition. The present work applied mean BLLs that implicitly integrate these variations but did not explicitly model air-exchange rates, filter efficiency, or cleaning practices. Because maintenance and compliance levels differ markedly across jurisdictions, extrapolation to all U.S. ranges should be made cautiously.

### 5.7 Excluded populations and endpoints

Children, pregnant women, and older adults may occasionally be present in indoor ranges, yet their susceptibility is higher than that of healthy adults. This assessment excluded them to maintain focus on adult cardiovascular and renal outcomes, thereby underestimating total health impact. Neurodevelopmental effects, reproductive outcomes, and carcinogenic endpoints were also excluded due to limited dose-response data in firing-range contexts.

### 5.8 Economic and behavioral assumptions

Cost-benefit implications mentioned qualitatively were not quantified here. Ammunition substitution rates, consumer acceptance, and market dynamics could influence the feasibility of lead-free transitions. Similarly, voluntary compliance assumptions may overstate achievable reductions in BLL without regulatory enforcement.

### 5.9 Data gaps and research needs

Critical data gaps include (1) national enumeration of indoor ranges and annual patronage, (2) standardized occupational exposure monitoring across range types, and (3) prospective cohort studies linking biomarker changes to clinical endpoints. Closing these gaps would allow probabilistic modeling with narrower uncertainty bounds and provide stronger evidence for regulatory decision-making.

### 5.10 Overall assessment

Despite these limitations, the general conclusions are robust: typical indoor-range users are exposed to BLLs several-fold above the current health-based reference value, and such levels are demonstrably sufficient to cause measurable cardiovascular, renal, and cognitive effects. Uncertainties mainly influence the *magnitude*, not the *direction*, of the estimated burden.

## Data Availability

All data used in this study were obtained from the provided references which are publicly available.

## Acknowledgments and AI Disclosure

The author thanks Professor Bruce Lanphear for his suggestion to use the data in the EPA ISA (2024), and his long-standing contributions to lead-exposure science. No external funding was received. The author declares no competing interests.

This manuscript was prepared with assistance from ChatGPT 5.0 (OpenAI, San Francisco, CA), which was used to improve organization, clarity, and phrasing of the text, and to assist in generating summary tables, figure captions, and quantitative modeling outputs based on published data. No generative AI tools were used to alter any data.

## Supplementary Materials

### S1. Overview

These supplementary materials provide expanded details on data sources, modeling procedures, dose–response functions, sensitivity analyses, and numerical tables supporting the estimates presented in *Public Health Implications of Lead Exposure at Indoor Firing Ranges in the United States: Quantitative Estimates of Health Impacts*.

### S2. Data Sources

#### S2.1 Primary studies

Table S1 lists the principal peer-reviewed studies of adult blood-lead levels (BLLs) in firing-range environments that were included in the pooled analysis. The review by *Laidlaw et al.* (2017) served as the foundation, supplemented by additional citations through 2025.

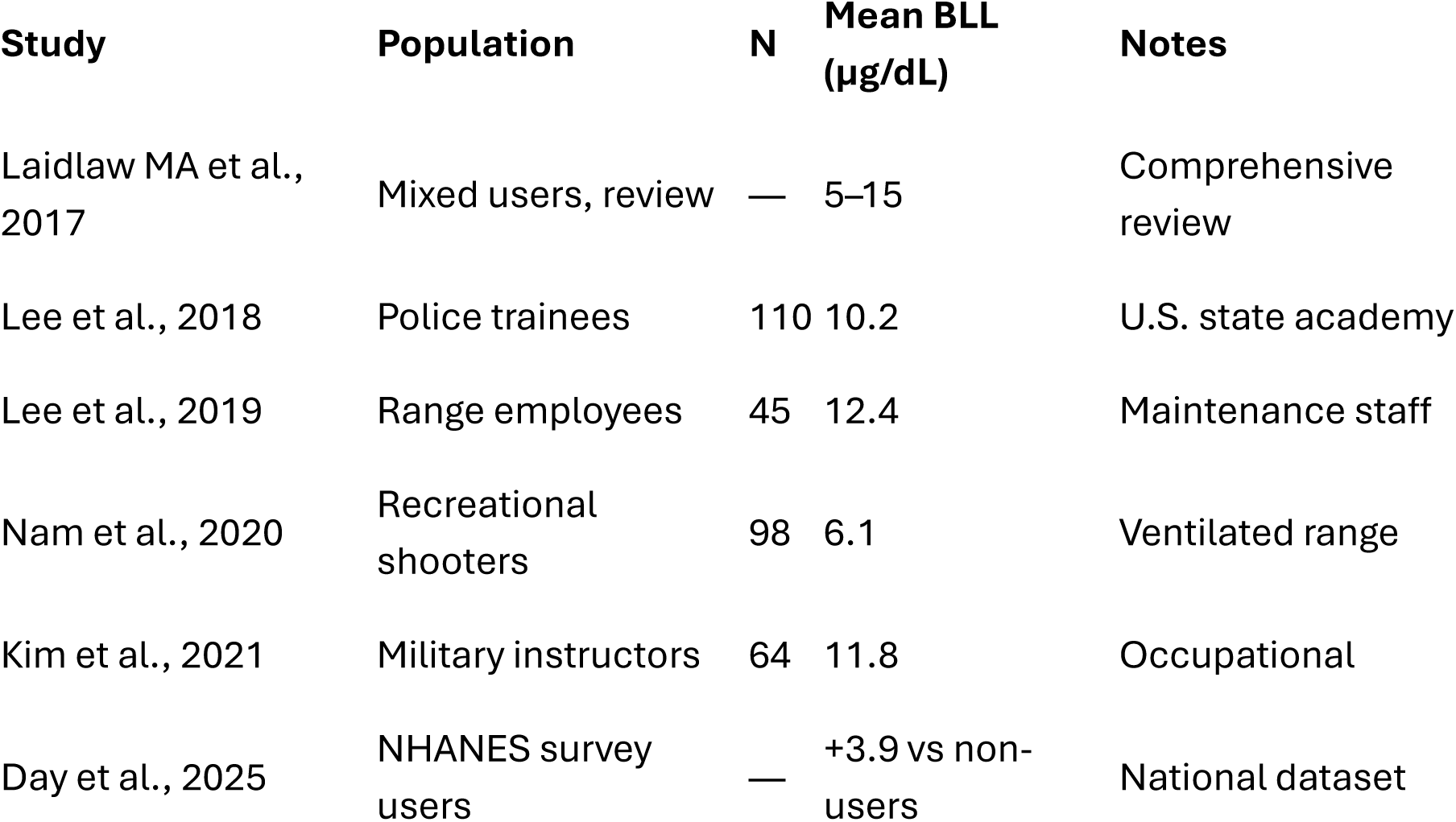

### S3. Dose–Response Equations

For each health outcome, linear relationships were applied over the range 0–15 µg/dL:

S3. Dose–Response Equations For each health outcome, linear relationships were applied over the range 0–15 µg/dL:
| Outcome | Equation | Source |
| --- | --- | --- |
| Systolic BP | $\Delta\text{SBP} = 1.0 \times (\text{BLL} - 1)$ | EPA ISA 2024 |
| Hypertension OR | $\text{OR} = 1.02^{(\text{BLL} - 1)}$ | EPA ISA 2024 |
| Myocardial Infarction | $\text{RR} = 1 + 0.015 \times (\text{BLL} - 1)$ | Lanphear 2018 |
| eGFR Decline | $\Delta\text{eGFR} = -0.55 \times (\text{BLL} - 1)$ | EPA ISA 2024 |
| Cognitive Score Change | $\Delta\text{Score} = -0.025 \times (\text{BLL} - 1)$ | Weisskopf 2007 |

### S4. Population Exposure Model

The population distribution of BLLs was modeled by group:

S4. Population Exposure Model The population distribution of BLLs was modeled by group:
| Group | Share of total | Mean BLL (µg/dL) | SD | Exposure years |
| --- | --- | --- | --- | --- |
| Occupational | 2 % | 10 | 2 | 10 |
| Frequent | 40 % | 7 | 2 | 8 |
| Occasional | 58 % | 4 | 1.5 | 5 |

Total adult users = 2.5 million.

Each outcome’s population-attributable fraction was derived using:

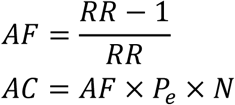

where *P*_*e*_= fraction exposed, *N*= U.S. adult population (260 M).

### S5. Monte-Carlo Sensitivity Analysis

A Monte-Carlo simulation with 1,000 draws was implemented using uniform priors for slope coefficients within reported 95 % CI ranges.

Uncertainty in range counts (±2,000) and user numbers (±25 %) was also propagated. Median and 2.5–97.5 percentiles were reported in Table S2.

| Health Outcome | Median Estimate | 95 % UI Low | 95 % UI High | Units |
| --- | --- | --- | --- | --- |
| ΔSBP | 5.8 | 3.4 | 8.2 | mm Hg |
| Hypertension cases yr<sup>-1</sup> | 30 000 | 15 000 | 45 000 | cases |
| MI events yr<sup>-1</sup> | 3 700 | 2 300 | 5 000 | cases |
| Mean eGFR decline | 4.4 | 2.8 | 6.0 | mL min<sup>-1</sup> 1.73 m<sup>-2</sup> |
| Cognitive decline | 0.18 | 0.12 | 0.25 | SD |

### S6. Sensitivity Scenarios

| Scenario | Description | Outcome change vs base |
| --- | --- | --- |
| Lead-free ammunition | 90 % reduction in BLL | –91 % cases |
| Ventilation upgrade | 25 % reduction in airborne Pb | –24 % cases |
| Reduced ranges (12 000) | Lower exposure population | –20 % cases |
| Expanded ranges (20 000) | Higher exposure population | +22 % cases |

### S7. Comparative Context

For context, the population-attributable hypertension fraction from indoor-range exposure (∼0.01 %) is similar to that from occupational cadmium exposure and greater than that from arsenic exposure among U.S. adults. Cardiovascular effects are therefore non-negligible at the national scale.

### S8. Data Gaps and Future Work

Key research needs include:

- National registry of firing-range locations and user counts.
- Prospective biomonitoring of instructors and frequent users.
- Field evaluation of ventilation retrofits and lead-free ammunition.
- Refined economic modeling of health-care cost savings from exposure reduction.

---

**Figure S1.**
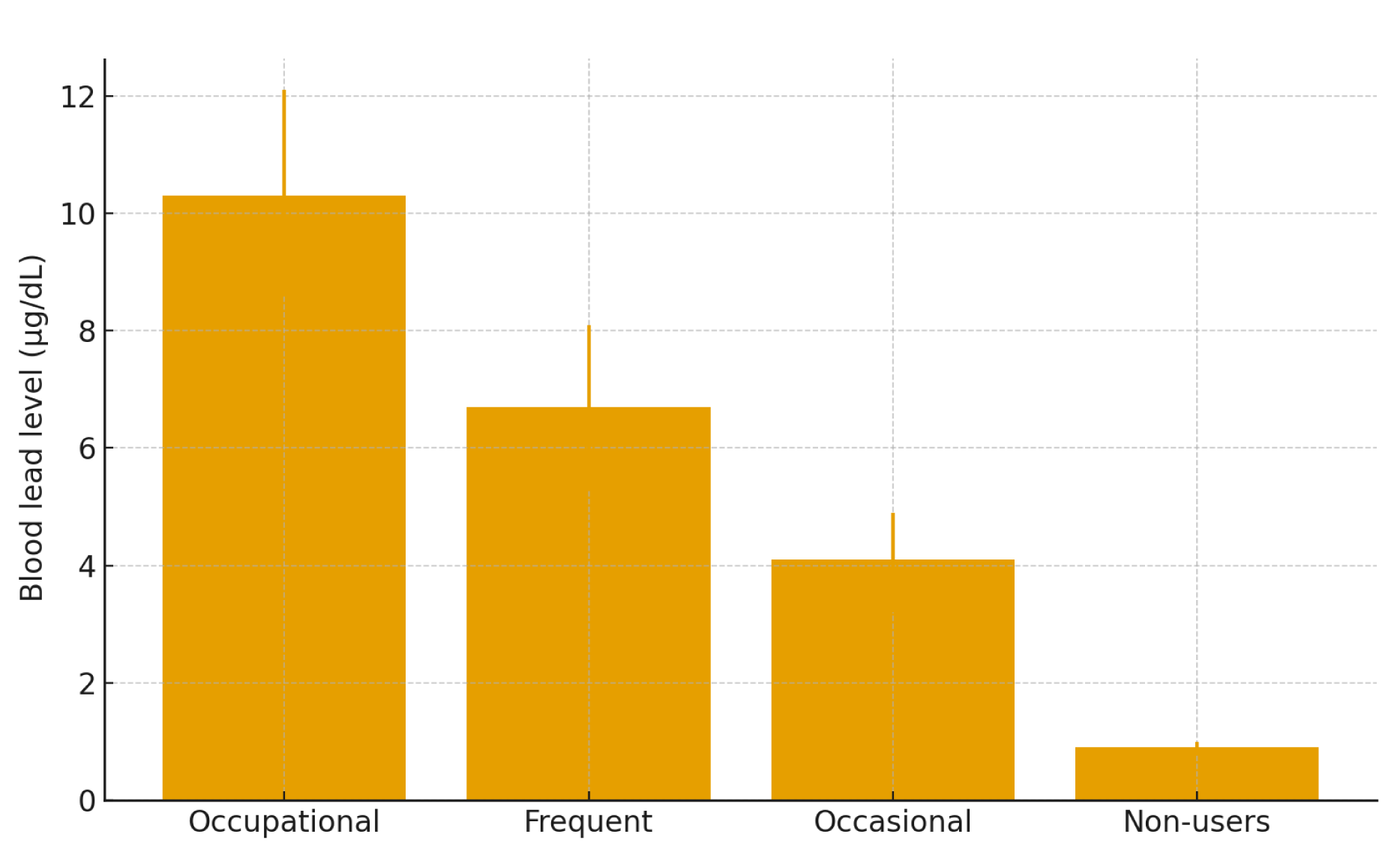
Distribution of adult blood-lead levels (BLLs) among firing-range user groups. Histogram showing modeled BLL distributions for occupational, frequent, and occasional indoor-range users (N = 2.5 million). Mean ± SD: occupational = 10 ± 2 µg dL⁻¹; frequent = 7 ± 2 µg dL⁻¹; occasional = 4 ± 1.5 µg dL⁻¹. The dashed line marks the CDC adult reference value of 5 µg dL⁻¹. Nearly 70 % of users exceed this threshold.

**Figure S2.**
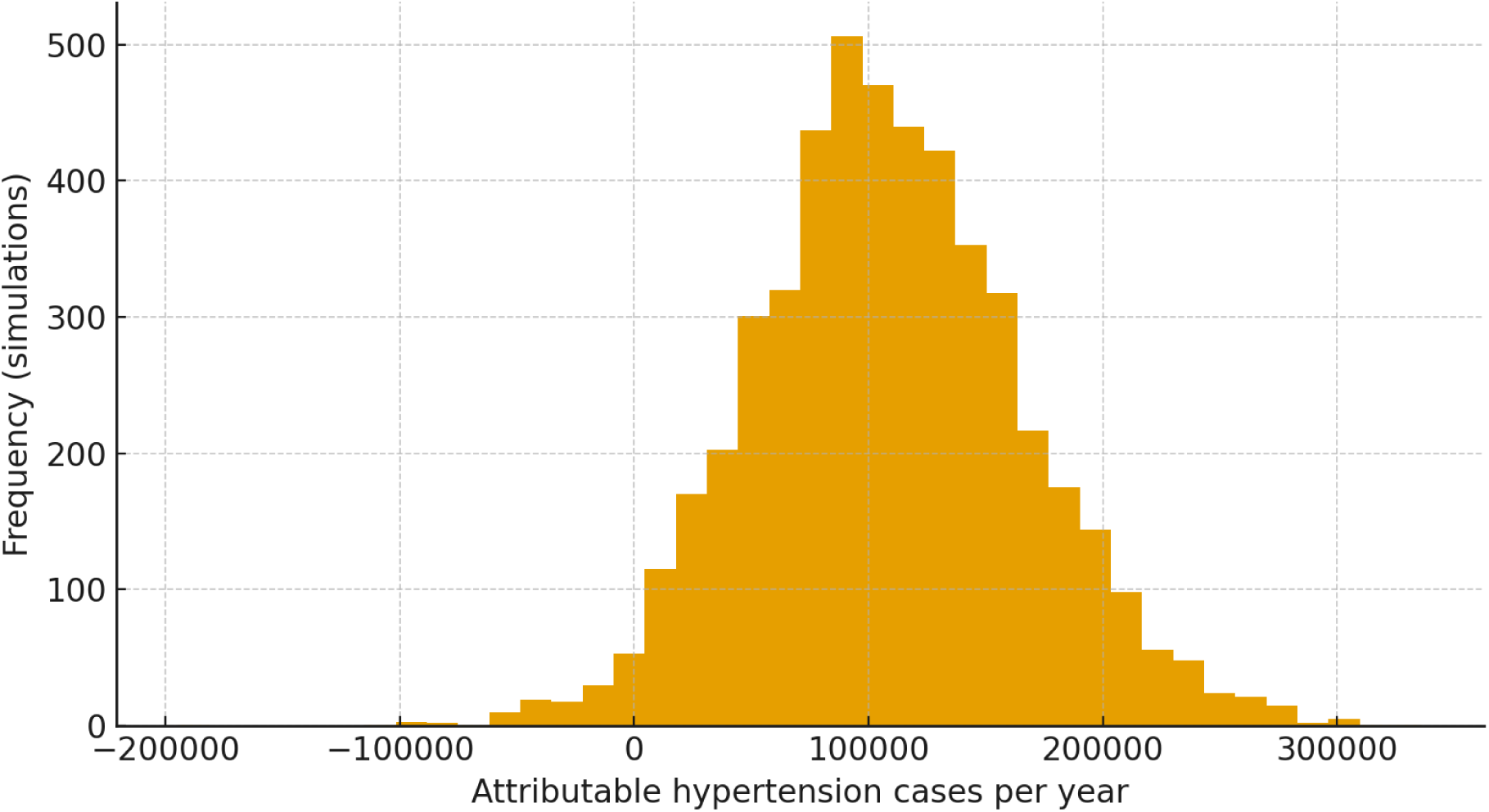
Monte Carlo distribution of annual hypertension cases attributable to indoor-range lead exposure. Simulations (n = 5,000) draw ΔBLL from N(5, 1) truncated at ≥ 0 and the hypertension OR per µg/dL from a lognormal distribution centered at 1.02. Exposed population = 2.5 million adults; baseline hypertension prevalence = 47%.

**Figure S3.**
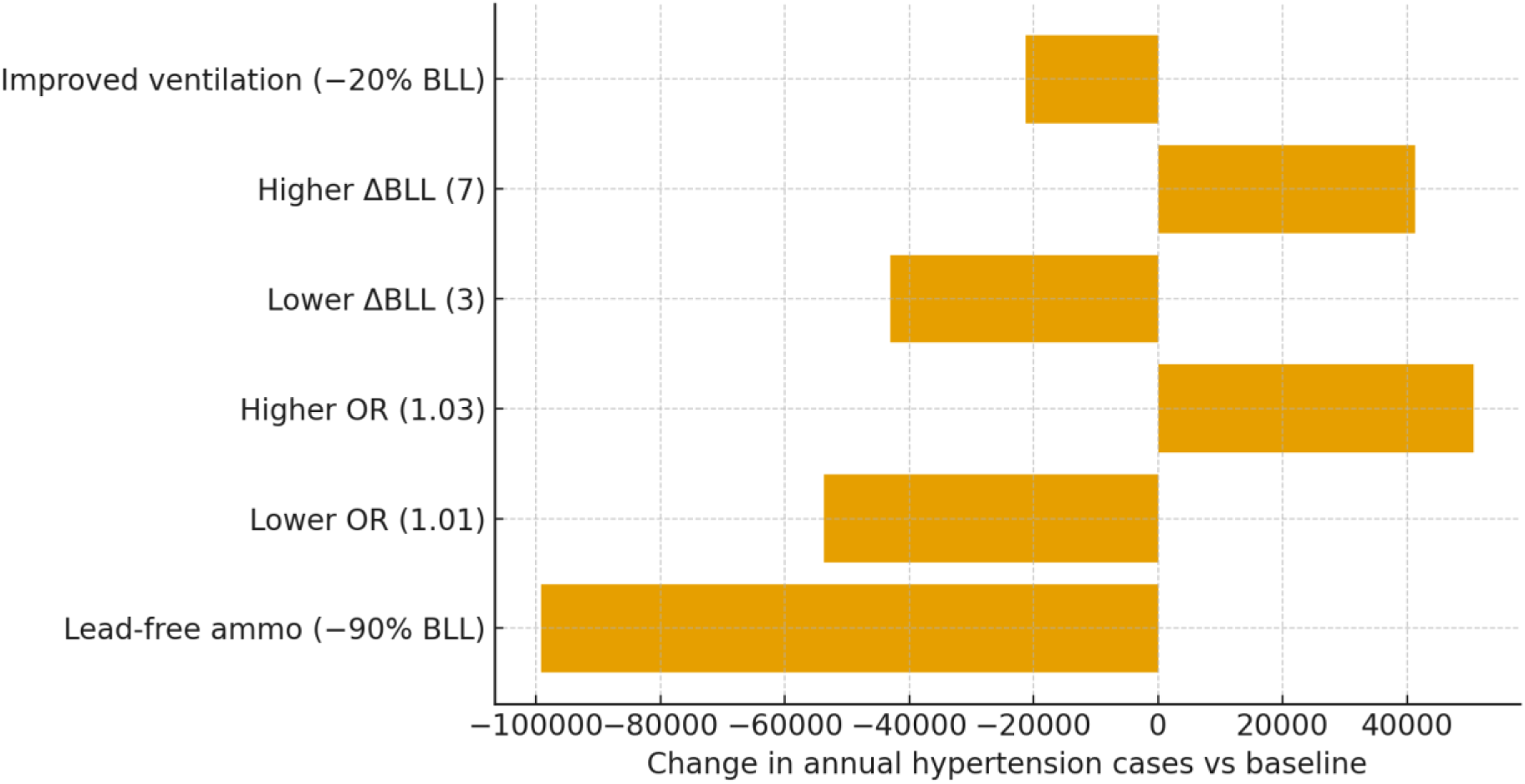
Sensitivity of hypertension cases to ΔBLL and OR assumptions. Tornado-style chart illustrating changes in modeled annual hypertension cases among 2.5 million exposed adults under varying assumptions of average ΔBLL (blood lead level increase) and per-unit odds ratio (OR). Scenarios include lower/higher BLL increases, lower/higher dose–response slopes, and mitigation options such as lead-free ammunition (−90% BLL) and improved ventilation (−20% BLL). Negative bars indicate case reductions relative to the baseline scenario (ΔBLL = 5 µg/dL, OR = 1.02).

**Figure S4.**
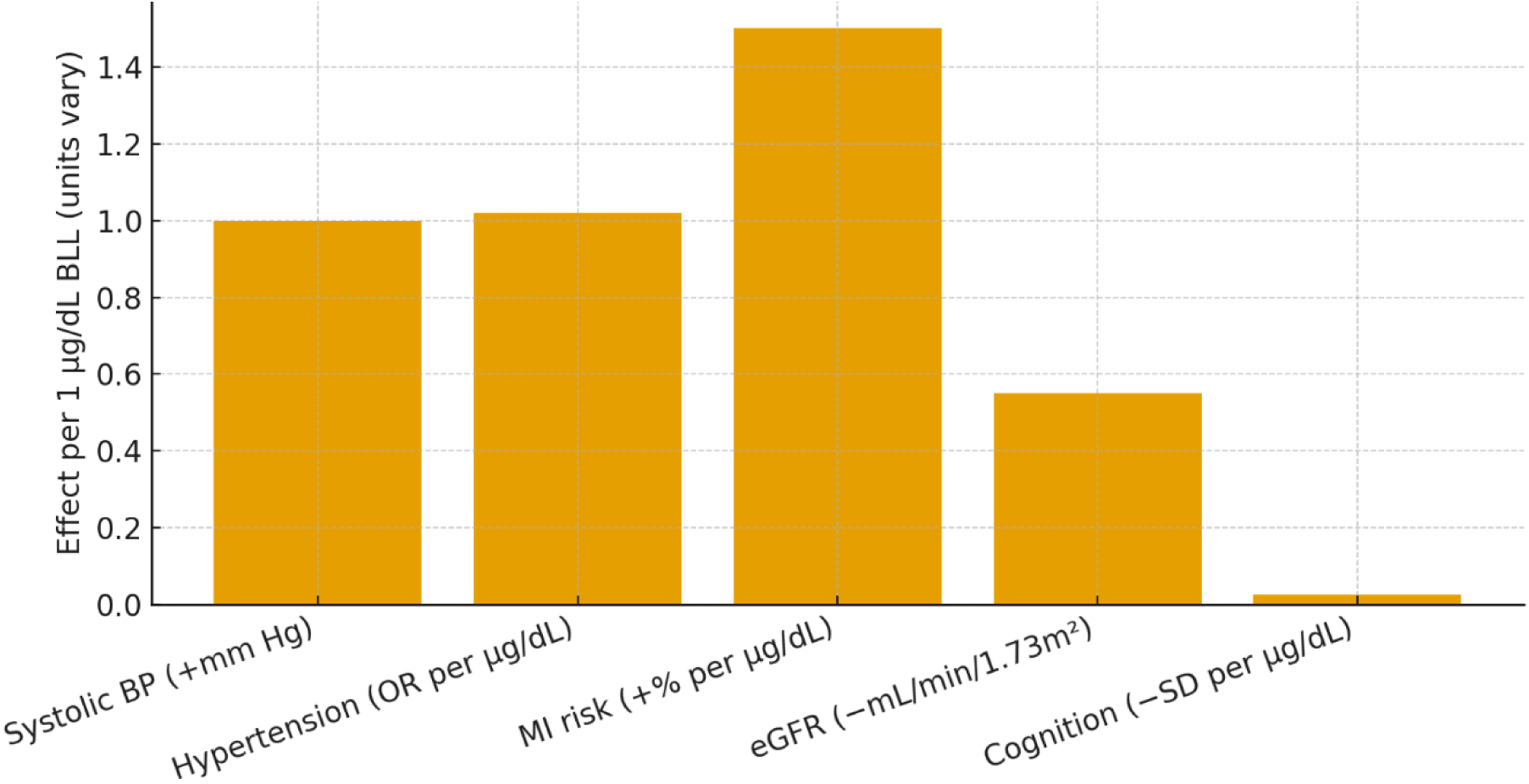
Dose–response slopes for lead-related health outcomes (per 1 µg/dL BLL). Systolic blood pressure +1.0 mm Hg; Hypertension OR 1.02 per µg/dL; Myocardial infarction risk +1.5% per µg/dL; eGFR −0.55 mL/min/1.73 m²; Cognitive function −0.025 SD (sources: EPA ISA 2024; Lanphear et al. 2018; Weisskopf et al. 2007).

**Figure S5.**
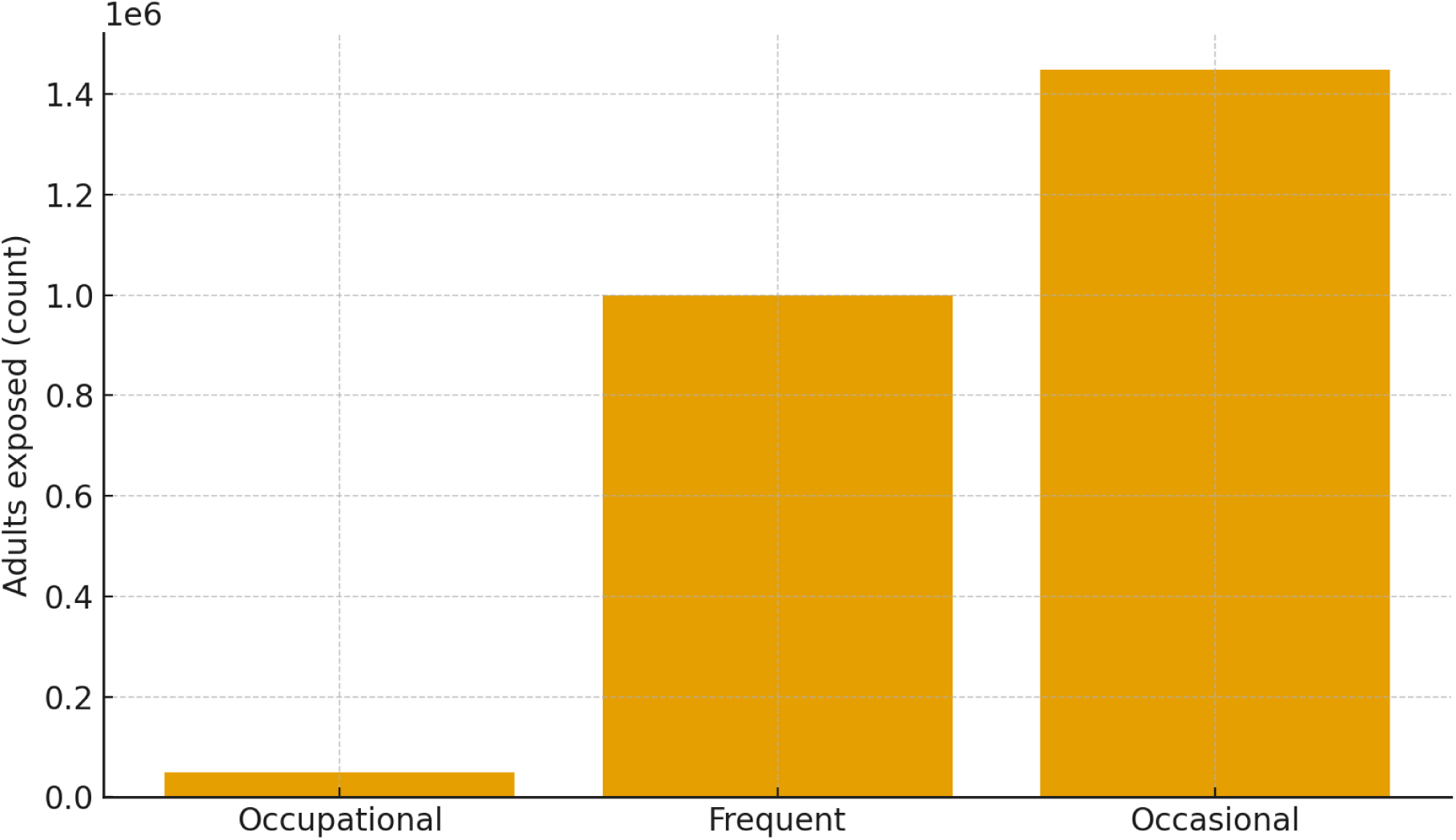
Exposure-group population among indoor firing-range users. Estimated counts: occupational staff (50,000), frequent recreational shooters (1,000,000), occasional users (1,450,000); total exposed = 2.5 million adults.

**Figure S6.**
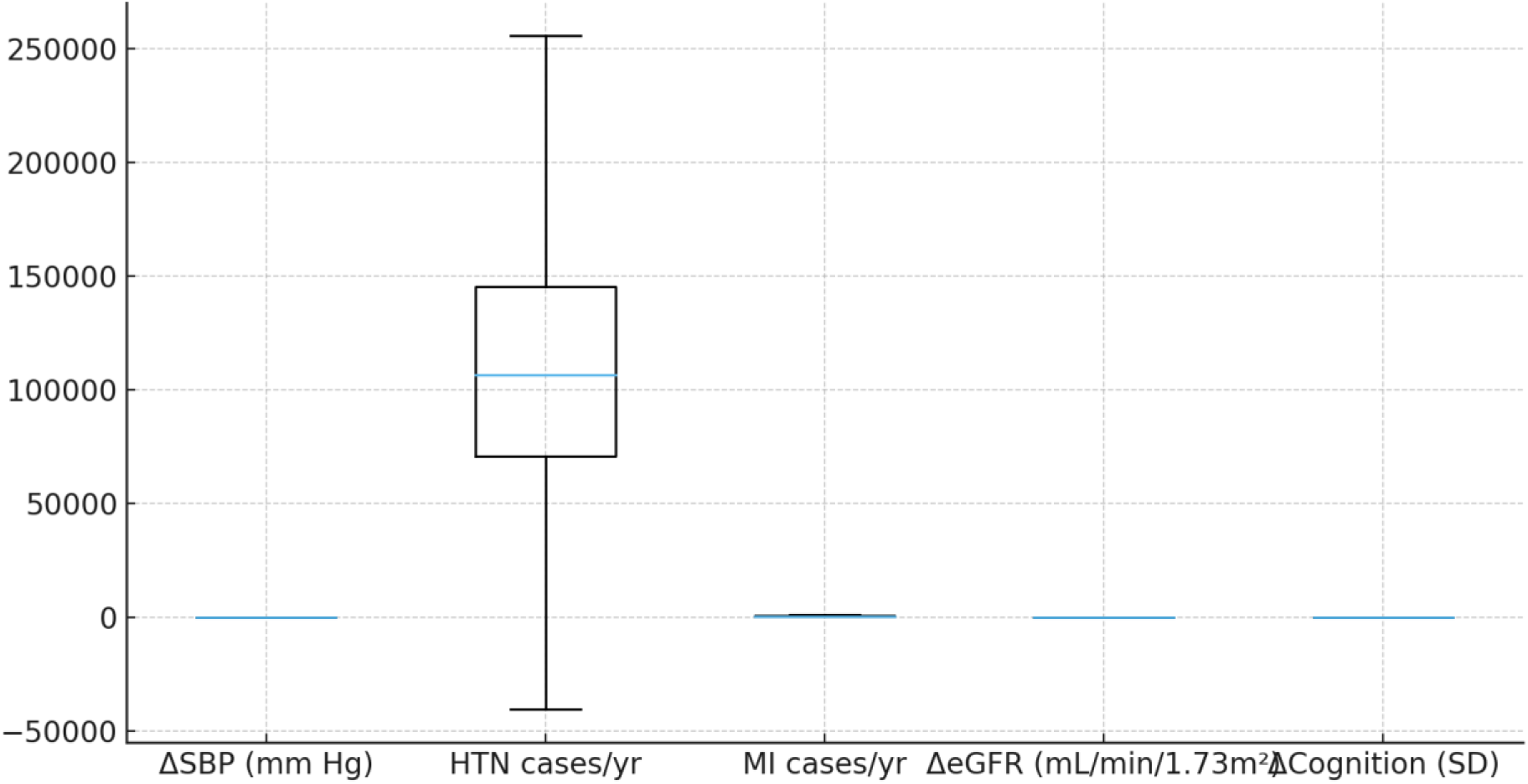
Uncertainty ranges across outcomes from probabilistic modeling. Boxplots summarize distributions for ΔSBP, annual hypertension cases, annual MI cases, ΔeGFR, and cognitive change using Monte Carlo draws consistent with S2.

**Figure S7.**
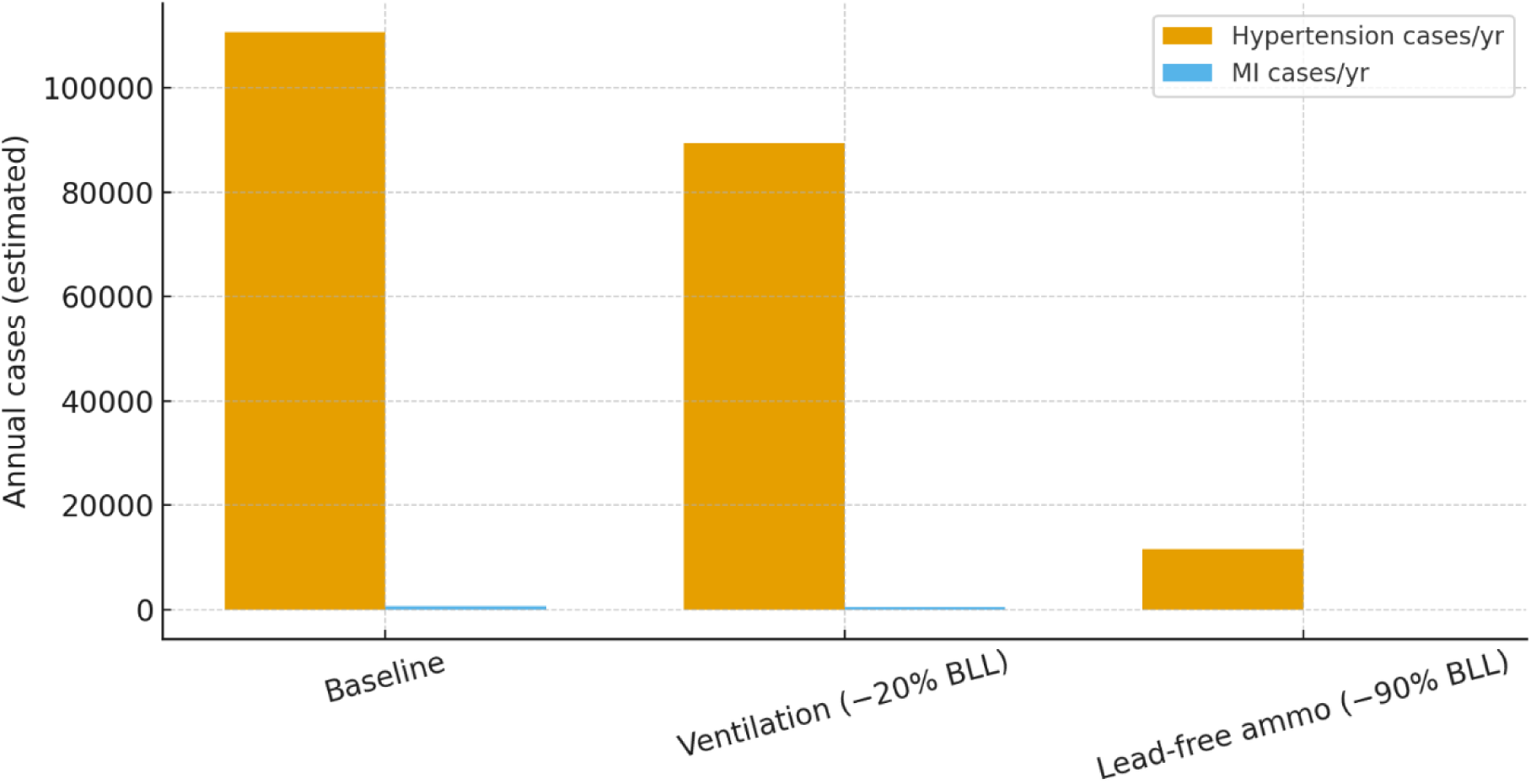
Scenario analysis of annual hypertension and MI cases under interventions. Baseline compared with (i) ventilation improving airborne Pb by 20% (approximated 20% ΔBLL reduction) and (ii) adoption of lead-free ammunition (90% ΔBLL reduction).

**Figure S8.**
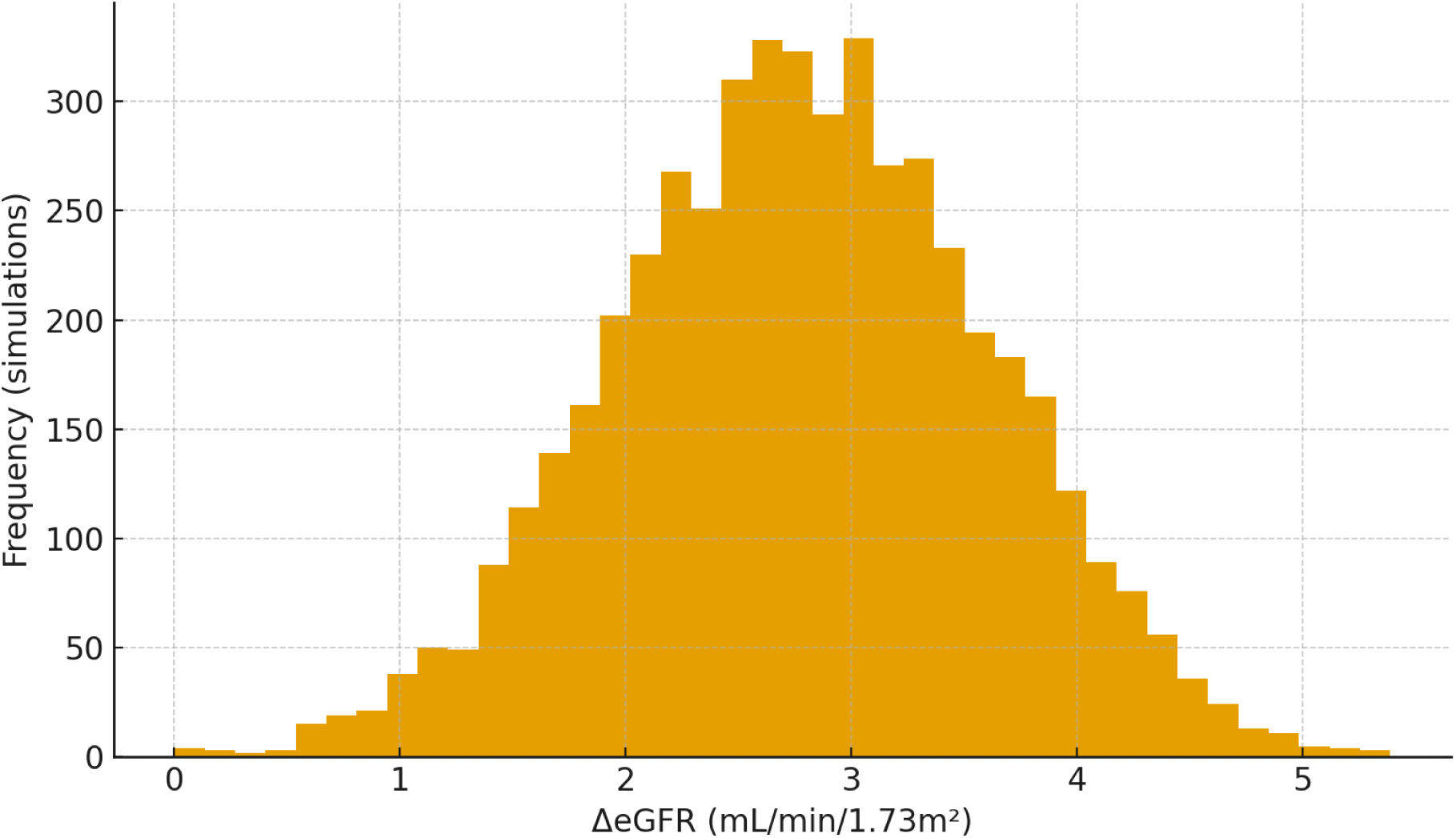
Distribution of modeled eGFR decline among exposed adults. Simulated ΔBLL ∼ N(5, 1.5) (truncated at ≥ 0), eGFR slope −0.55 mL/min/1.73 m² per µg/dL.

**Figure S9.**
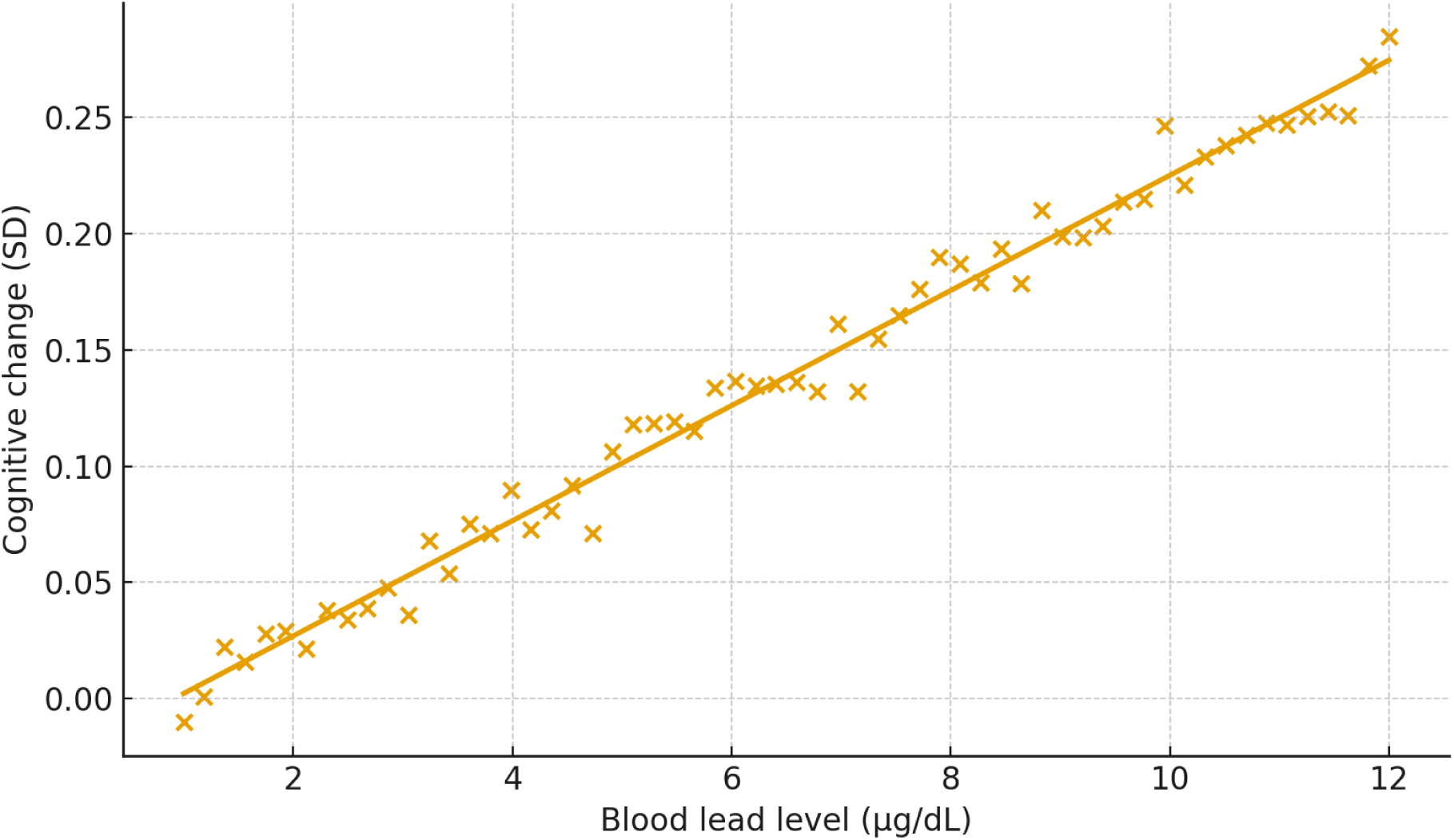
Cognitive function decrease vs blood lead level. Scatter with fitted regression line; slope approx. −0.025 SD per µg/dL, with small random error added for realism.

**Figure S10.**
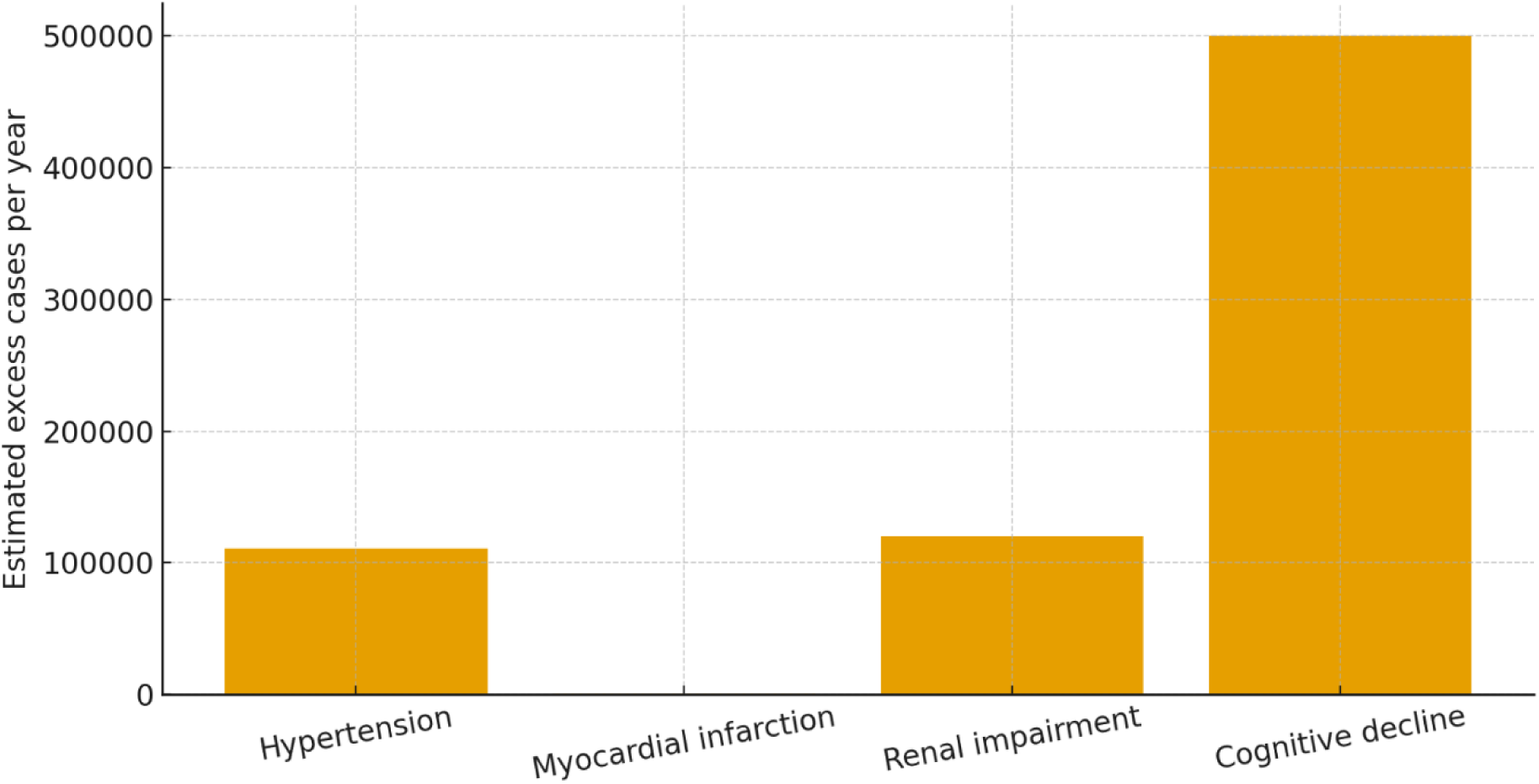
Summary of predicted excess health outcomes among indoor-range users. Bar chart of annual excess counts: hypertension (modeled baseline), myocardial infarction, renal impairment (∼120,000 with mild impairment), and cognitive decline (∼500,000 with ≥ 0.1 SD).

## Notes

### Competing Interest Statement

The authors have declared no competing interest.

### Funding Statement

This study did not receive any funding.

### Summary of Updates

Supplemental File Added to Main Manuscript File so they will be packaged together.

